# A comparative national-level analysis of government food system resilience activities in preparation for future food system disruptions

**DOI:** 10.1101/2023.04.19.23288822

**Authors:** Jane Lloyd, E.R.H. Moore, Lyndsey Dowell, Roni Neff

## Abstract

**Background:** The COVID-19 pandemic, extreme weather events, and the Russian invasion of Ukraine have highlighted global food system vulnerabilities and a lack of preparedness and prospective planning for increasingly complex disruptions. This has spurred an interest in food system resilience. Despite the elevated interest in food system resilience, there is a lack of comparative analyses of national-level food system resilience efforts. An improved understanding of the food system resilience landscape can support and inform future policies, programs, and planning.

**Methods:** We conducted a cross-country comparison of national-level food system resilience activities from Australia, Aotearoa (New Zealand), Sweden, and the United States. We developed upon and adapted the resilience framework proposed by Harris and Spiegel to compare actions derived from thirteen national food system resilience documents. We coded the documents based on how the governments determined actions by food system resilience attribute utilized, part of the food supply chain, specific shocks or stressors, implementation level, the temporal focus of action, and the expected impact on food security. We analyzed and compared countries’ coded categories, subcategories, and category combinations.

**Results:** The results showed that countries are using multi-pronged policy actions to address food system resilience issues and are focused on both retrospective reviews and prospective models of disruptive events to inform their decisions. Some work has been done towards preparing for climate change and other natural disasters, but not as much for other shocks or stressors.

**Conclusions:** The analysis identified potential gaps, concentrations, and themes in national food systems resilience. The framework can be applied to augment existing policy, create new policy, as well as to supplement and complement other existing frameworks.

## Background

Our global food systems are at risk from natural and human-made disasters. The Coronavirus Disease 19 (COVID-19) pandemic highlighted a concatenation of food systems issues [1] at national levels due to a lack of preparedness and recognition of existing vulnerabilities [2] and lack of foresight and prospective planning for new and more complex shocks [3]. The direct impacts of COVID-19 were felt within and across countries’ food systems, requiring governments and societies to respond – globally, nationally, sub-nationally, and at the community and household level. The Russian invasion of Ukraine had a compounding effect and prompted an international response: the United Nations created a “Global Crisis Response Group on Food, Energy, and Finance” to support policymakers in mobilizing solutions and developing strategies to address the impact of rising energy prices on the cost-of-living crisis, food insecurity, and social unrest [4].

Even without these crises, global food and social systems have been failing to meet the nutritional adequacy requirements of many populations. Food-related non-communicable disease has risen and is now the leading cause of death globally [5]. Food insecurity and undernutrition (malnutrition and obesity) are prevalent in low-, middle- and high-income countries. These nutritional challenges are often exacerbated by food system disruptions, such as droughts causing a decrease in crop growth and resulting in famine, with impacts disproportionately harming populations at risk of or facing food insecurity [6].

As a result of these compound events, there has been increased interest at global, national, and subnational levels in food system resilience. A resilient food system, as defined by the Johns Hopkins Center for a Livable Future (adapted from Tendall et al. [7]), is “one that is able to withstand and recover from disruptions in a way that ensures a sufficient supply of acceptable and accessible food for all” [8]. A resilient food system is one with the ability to respond to and recover from disruptions that are either shocks (transitory adverse events) or stressors (persistent adverse trends) [9,10] having either natural or man-made origins. A shock, for example, could be an immediate natural disaster such as a hurricane that disrupts food production systems and access to food by destroying crops or roads, thus preventing food from reaching consumers. Stressors include long-term trends such as drought or desertification [9,10], declining resources such as declining fish stocks due to overfishing, or ongoing cybersecurity threats. A country’s level of food security can be used as one benchmark for its food system resilience [10], although the concept is far more complex. Food Security Information Network (FSIN), in the Resilience Measurement Principles [9], outlined that a country’s response to shocks and stressors should result in a household or community returning to a “normative” state, determined as “acceptable levels of well-being” [9]. Based on this definition, a food insecure community impacted by a shock should not returned to a food insecure state during the recovery phase of a shock, as the normative threshold is not being food insecure [10] but rather entering a new and better state. Candy et al. [11] notes that while food security is dependent on a food system’s being resilient, it is not indicative of a resilient food system.

Despite the increased interest in food system resilience, few governments at the national and local levels have conducted food system resilience reviews or policy planning. There are exceptions, for example: national-level – United States, completed by the United States Department of Agriculture (USDA) [12]; subnational – Maryland, United States [13]; municipal – Baltimore, Maryland, United States [14], Boston, Massachusetts, United States [15], Toronto, Canada [16], and Christchurch, New Zealand [17]. At local levels, several city councils, such as that of Auckland, New Zealand have advocated for food system resilience policies at a national level [18]. To our knowledge no prior reviews have compared national-level food system resilience planning documents.

To enhance the understanding of food system resilience policy landscapes, we conducted a comparative analysis of food system resilience planning documents by national governments for four countries (Australia, Aotearoa (New Zealand), Sweden, and the United States). We wanted to understand what countries viewed as their food system resilience concerns, how they addressed them, and how the countries compared to each other. By analyzing countries’ varied approaches to food system resilience, we aimed to identify approaches to inform policymaking in the future.

## Method

### Country and document selection

To date there is no central repository for government food system resilience plans. We selected government documents for analysis after completing internet searches for national-level food system resilience plans using the following search terms in different variations: food system, food system resilience, government, national, plan, planning, and resilience. From the results, we then selected four countries to be a part of the comparative analysis: Australia, Aotearoa (New Zealand), Sweden, and the United States. The four are considered peer nations as (a) they have established primary sectors (agriculture, forestry, fisheries and aquaculture) and are categorized by the World Bank as “high-income economies”; (b) they are geographically dispersed (North America, Europe, and Oceania) and have varied risk profiles; and (c) they have differing degrees of food insecurity [19]. Australia, Sweden, and the United States have published national-level food system resilience documents (reviews, policies, or strategies) in the last ten years and have taken different approaches to food system resilience. We included New Zealand due to its risk profile, its economic and trade dependence on its primary sector, the impact of recent extreme weather events, and to understand policy opportunities for the country to address food system resilience (see supplementary information file 4).

For each of the four selected countries we then reviewed their government websites and grey literature. Documents were selected based on whether they contained food system resilience in the title, mentioned food system resilience as a goal, or had an independent document section dedicated to food system resilience. A member of the research team then reviewed the documents. As New Zealand did not have a national food system resilience plan, we compiled a list of related plans for climate change and resilience across civil defense, health, primary industries, and indigenous peoples.

### Conceptual Model

Currently, there are only a few analysis frameworks for policymakers to assess food system resilience. To assess the countries’ national-level food system resilience activity, we adapted the Harris et al. [10] framework as it provided a more complete set of food system resilience attributes outlined in the literature, while making the link between food system resilience policy attributes and the intended effect on food security.

We adapted the framework in several ways, based on updates in the literature [8,20,34] and deductive review of the data. We expanded the framework to include additional food system resilience attributes (adaption, awareness, capital reserves, connectivity, diversity, equity, redundancy, and preparedness) based on work done by members of the research team, and that are established in the literature [8], removed self-regulation (due to the inability to determine food system self-regulation from national-level documents), and extended the definition of food security to include sustainability and agency, to align with the Committee on World Food Security High Level Panel of Experts [20]. We added the following categories that were originally noted by Harris et al. [10], but not used in their analysis: part of the food supply chain (producer, processor, distributor, input services, and support services) [34] where the action is being targeted; the shock or stressor related to the issue or action being addressed; the implementation level to which actions were directed – national, regional, local/state, community or household; and temporal focus, including an assessment of when actions are required in response to the effects of shocks and stressors—in the short, medium, or long term. We also added a component indicating whether the actions were taken to a shock or stressor from a prospective or retrospective perspective. We provide this adapted conceptional framework in Figure 1, that was used as a guide for the analysis.

**Fig. 1.**
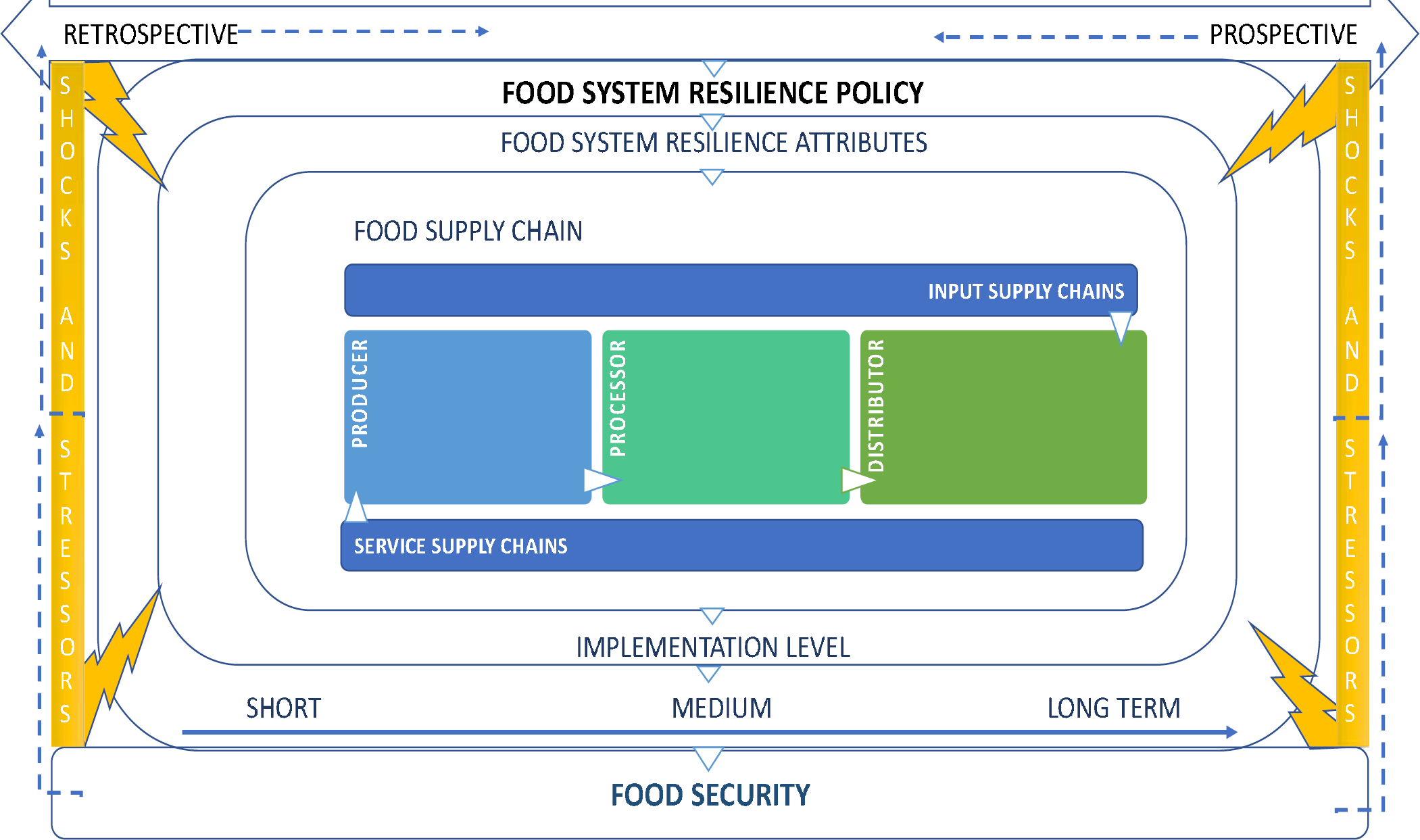
Conceptual framework for analysis. A country’s food system resilience is reviewed using retrospective and/or prospective data from shocks and stressors; these previous or expected disruptions inform food system resilience issues and actions. Food system resilience policy is developed and includes actions that represent food system resilience attributes and part of the food supply chain, and are implemented at different levels in society within a designated timeframe. The resulting actions have an expected impact on food security.

### Data Collection and Coding

For each national document included in the study, we reviewed the content and classified it into food system resilience “issues” and “actions.” Issues are specific concerns raised by or commissioned by the government that highlight where the country has determined that it was not or is not sufficiently prepared or equipped to maintain its food system. Actions are government-determined activities either undertaken or required to be undertaken in the future to address the issues raised to increase food system resilience. For example: the US document highlighted the issue of concentration and consolidation in agri-food production, manufacturing, and distribution, which they propose to address with the action of investing $4 billion in building regional and local facilities [11]. We coded the issue as related to concentration and consolidation in the food supply chain and the action as investing in local and regional alternative infrastructure.

We coded each issue or action by the seven categories and thirty-eight sub-categories outlined in our conceptual framework (Figure 1) and Table 2. We provide a full description of the categories and sub-categories and the source in supplementary information file 1, but, in brief: (1) Food system resilience attributes (adaption, awareness, capital reserves, connectivity, diversity, equity, redundancy, preparedness) are the characteristics that have been identified to enhance resilience in the food system by absorbing and/or mitigating the effects of disruptions [8]; (2) Part of the food supply chain or the main constituent components that enable the flow of food from production to consumption; (3) Anticipated stressors and shocks that can cause disruption to the food system; (4) Level of society at which the government actions will be implemented; (5) Expected impact of the food system resilience actions on food security; (6) Designated timeline for an issue or action to take place; and (7) Perspective from which the issue or action is derived—retrospectively when a disruptive event or events have already occurred, or prospectively when a disruptive event is predicted to occur.

The coding was done by a member of the research team (JL) and then reviewed for accuracy (JL). Where categorizations could be interpreted in various ways, the assigned codes were discussed and agreed within the research team (JL, EM) prior to the results being finalized. A random check for coding accuracy was done by another member of the research team (EM). For each action, at least one sub-category needed to be selected per category. Coding was done as 1 or 0, with 1 indicating the that the document referenced the subcategory and 0 indicating no reference. Three exceptions to that approach were: when no timeline was stipulated that category was left blank; when a specific subcategory of shock or stressor was not stipulated, all subcategories were coded with a 1; and in category 1 capital reserves and other were both coded with a 1 when “funding” needs were outlined, as funding does not fit neatly into the definition of capital reserves.

### Data analysis

To determine countries’ breadth and depth of actions, we calculated frequencies of actions by category and sub-category. We then calculated the percentage that each sub-category comprised of a country’s total actions within that resilience category (Table 2 and supplementary information file 2 and 3). We also assessed the extent to which multiple documents within a country repeated the same combinations across categories versus distinctive ones. We did this by distilling the combination frequencies of all policy actions and removing timeframe and perspective (Table 3 and supplementary information file 2).

## Results

Table 1 lists the documents included in this comparative analysis. We identified two documents from Australia, nine documents from New Zealand, one document from Sweden, and one document from the US. Some countries took a more consolidated approach with one comprehensive document, whereas others had multiple documents containing relevant food system resilience information.

**Table 1.** Thirteen national documents included in the comparative analysis.

| Document | Country |
| --- | --- |
| Resilience in Australian food supply chain (2012) [21] | Australia |
| Security of Critical Infrastructure Act 2018 [22] | Australia |
| Te hau mārohi ki anamata: Towards a productive, sustainable and inclusive economy: Aotearoa New Zealand’s First Emissions Reduction Plan (2022) [23] | New Zealand |
| Considerations for developing a Health National Adaptation Plan for New Zealand (2019) [24] | New Zealand |
| Exploring An Indigenous Worldview Framework for the National Climate Change Adaptation Plan (2021) [25] | New Zealand |
| Urutau, ka taurikura: Kia tū pakari a Aotearoa i ngā huringa āhuarangi Adapt and thrive: Building a climate-resilient New Zealand Aotearoa New Zealand's First National Adaptation Plan (2022) [26] | New Zealand |
| Fit for a better world: accelerating our economic potential. Ministry for Primary Industries (2019) [27] | New Zealand |
| Sustainability and the health sector: A guide to getting started (2019) [28] | New Zealand |
| National Disaster Resilience Strategy: Rautaki ā-Motu Manawaroa Aituā (2019) [29] | New Zealand |
| National climate change risk assessment for New Zealand/Arotakenga Tūraru mō te Huringa Āhuarangi o Āotearoa: Main Report (2020) [30] | New Zealand |
| New Zealand Critical Lifelines Infrastructure: National Vulnerability Assessment 2020 Edition [31] | New Zealand |
| A National Food Strategy for Sweden: more jobs and sustainable growth throughout the country (2016) [32] | Sweden |
| USDA Agri-Food Supply Chain Assessments: Program and Policy Options for Strengthening Resilience (2021) [12] | United States of America |

Two documents were included for Australia. The first, *Resilience in the Australian Food Supply Chain (2012)* [21] was published in the wake of several natural disasters and based on the recognition that there was a growing likelihood of compounding or coinciding disasters, and also that, at the time, resilience within Australian supply chains was not well understood [21]. The focus of the report was to understand the impact of the disasters on Australian residents, and the ability of the food supply chain to regain its capacity in the event of a crisis or disaster [20]. An outcome of the report was the inclusion of food companies in the Security of Critical Infrastructure Act [22].

At the time of writing, New Zealand does not have a national food system resilience policy or strategy. It has, however, indirectly addressed some aspects of resilience in its climate change policies and strategies (see Table 1). For the review, we selected nine documents from New Zealand that addressed one or more shocks or stressors and included aspects of the food system.

We reviewed one document from Sweden, *A National Food Strategy for Sweden (2016)* [32]. The document aims to set the food system’s path to 2030, with a focus on strategically developing Sweden’s ability to establish stable and long-term resilience in the food supply chain, even in the face of systemic challenges that included low profitability and tough international competition, while addressing global challenges such as climate change and environmental problems [32].

We reviewed one document from the United States, the *Agri-Food Supply Chain Assessment: Program and Policy Options for Strengthening Resilience (2021)* [12]. This document resulted from vulnerabilities revealed during the COVID-19 pandemic that needed to be addressed over the short and longer term.

Table 2 summarizes the number of issues and actions identified for each country, by category and subcategory. We identified 33 issues for Australia, 31 issues for New Zealand, 3 issues for Sweden, and 14 issues for the United States. We identified 31 actions for Australia, 54 actions for New Zealand, 19 actions for Sweden, and 76 actions for the United States.

**Table 2.** Number of identified issues and actions by country for each food system resilience category and subcategory.

|  | Australia | New Zealand | Sweden | United States |
| --- | --- | --- | --- | --- |
| **Issues** | **33** | **31** | **3** | **14** |
| **Actions** | **31** | **54** | **19** | **76** |
| **Food System Resilience** |  |  |  |  |
| **Attribute** |  |  |  |  |
| Adaption | 6 | 15 | 3 | 1 |
| Awareness | 9 | 16 | 5 | 23 |
| Capital Reserves | 3 | 2 | 0 | 26 |
| Connectivity | 12 | 5 | 2 | 9 |
| Diversity | 3 | 7 | 8 | 14 |
| Equity | 0 | 13 | 1 | 6 |
| Preparedness | 18 | 10 | 2 | 15 |
| Redundancy | 6 | 9 | 6 | 4 |
| Other | 0 | 0 | 0 | 20 |
| **Food Supply Chain** |  |  |  |  |
| Producer | 7 | 31 | 17 | 37 |
| Processor | 10 | 3 | 3 | 9 |
| Distributor | 18 | 13 | 7 | 5 |
| Support Services | 20 | 17 | 2 | 31 |
| Input Services | 7 | 13 | 2 | 10 |
| **Shocks and Stressors** |  |  |  |  |
| Biosecurity | 2 | 5 | 7 | 30 |
| Climate | 2 | 49 | 15 | 35 |
| Cybersecurity | 2 | 2 | 6 | 7 |
| Economic & Political Crisis | 2 | 3 | 6 | 7 |
| Epidemic or Pandemic | 2 | 5 | 6 | 25 |
| Natural | 31 | 6 | 6 | 9 |
| Other | 0 | 0 | 0 | 20 |
| **Implementation Level** |  |  |  |  |
| National | 20 | 47 | 19 | 69 |
| Regional | 9 | 5 | 2 | 5 |
| Local | 19 | 18 | 2 | 10 |
| Community | 5 | 17 | 1 | 4 |
| Household | 5 | 0 | 0 | 1 |
| **Food Security** |  |  |  |  |
| Access | 3 | 9 | 2 | 1 |
| Agency | 0 | 9 | 2 | 3 |
| Availability | 27 | 18 | 13 | 34 |
| Stability | 2 | 3 | 0 | 14 |
| Sustainability | 1 | 31 | 9 | 39 |
| Utilization | 2 | 1 | 2 | 3 |

### Comparative Analysis of Food System Resilience Actions

Figure 2a compares the number and percentage of actions targeting each food system resilience attribute across countries. For Australia, most actions (58%) focused on preparedness. Sweden’s actions focused on increasing diversity (42%) and redundancy (32%) of the food supply to build resilience through increased production to manage its shortfalls between production and consumption. The United States had policy actions across all of the studied attributes, with capital reserves (34%) and “other” attributes (26%) showing the importance the United States is placing on funding to support existing and new resilience programs. New Zealand also outlined funding shortfalls in its need for infrastructure climate adaptation. The United States placed emphasis on awareness (30%) through expanding research and monitoring of food systems, in particular biosecurity due to climate change. New Zealand also placed an emphasis on developing awareness (30%) through monitoring the changes resulting from climate change that could impact agribusinesses. It also emphasized the use of equity (24%) by including the indigenous Māori worldview of climate change adaption, including specific references to the effects of climate change on Māori, their cultural and food gathering sites, and wellbeing.

**Fig. 2a-e.**
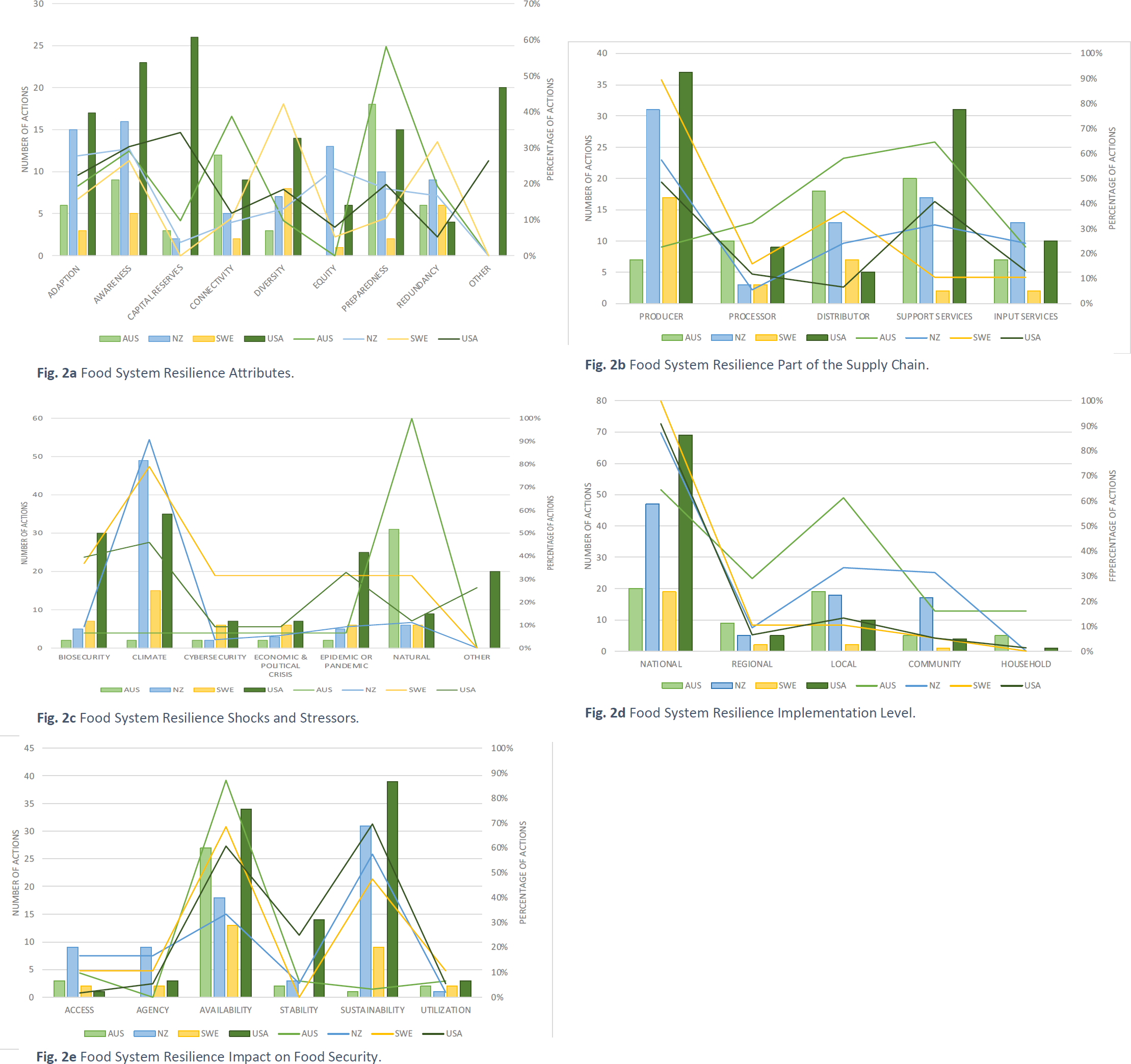
Comparison of Food System Resilience Actions Fig. 2a Food System Resilience Attributes. Adaption: having the food system flexible and able adapt to changing circumstances, modifying behaviors, and adapting existing resources to new purposes. Awareness: the food system has knowledge of its assets, liabilities, and vulnerabilities, including situational awareness. Capital Reserves: having social, financial, natural, political, food, and food input and supply reserves “backup” resources that can be used during a disruptive event. Connectivity: policies that promote integration and coordination among food system components. Diversity: having a variety of food system elements that can serve a similar purpose. Equity: having equity in food system resilience processes: procedurally, distributionally, structurally, and intergenerationally. Preparedness: having a plan in place for how to ensure food access, availability, acceptability, and agency during a disruptive event. Redundancy: having multiple or duplicative food system elements that can serve the same purpose. Fig. 2b Food System Resilience Part of the Supply Chain. Producer: the producer category includes food from agricultural and horticultural origins. Processor: a food processor means a food establishment that processes, manufactures, wholesales, packages, or labels food. Distributor: refers to a food retailer or food service provider. Support Services: include actors and activities for movement of inputs, outputs, and factors such as transport and storage operators, connecting production to consumption. Input Services: provide variable inputs, such as seed, fertilizer, fuel and labor, and quasi-fixed inputs, such as farm machinery, milling machines and coolers for perishables. Fig. 2c Food System Resilience Shocks and Stressors. Biosecurity: refers to harmful pests and diseases that can cause damage to plants and animals. Climate: refers to the stressor of climate change that has a multiplying effect to other stressors or shocks. It includes the effects of sea level rise, increased temperatures, coastal erosion, and more frequent extreme weather events. Cybersecurity: refers to shocks to digital technologies by exploited controls and practices to gain initial access or as part of other tactics to compromise cyber systems. Economic & Political Crisis: refers to a shock that is economic and political in nature (domestic or international in origin) that can have an unexpected large-scale impact on the economy. Epidemic or Pandemic: refers to a human disease outbreak that, in the case of an epidemic, has an unexpected increase in the number of disease cases in a specific geographical area and that, in the case of a pandemic, exhibits disease growth that is exponential and covers a wide area, affecting several countries and populations. Natural: refers to shocks or disasters that occur naturally, such as earthquakes, tsunamis, hurricanes/cyclones, tornados, landslides, floods, and droughts. Fig. 2d Food System Resilience Implementation Level. Fig. 2e Food System Resilience Effect on Food Security. Access: policies that make healthy food more financially and physically accessible. Agency: policies that consider an individual’s right to food, and fair and equal consideration of communities that affect the food system. Availability: policies that increase the amount of food in the food system. Stability: policies and planning that reduce instability or variability in the current food system from causes such as biosecurity crises. Sustainability: policies that reduce the impacts of on the future food system from causes such as degradation of natural resources. Utilization and Acceptability: policies that ensure food that is safe, acceptable, culturally appropriate, and provides sufficient nutrients and micronutrients to maintain good health.

Figure 2b compares the number and percentage of actions targeting each part of the food supply chain across countries. The comparison shows that all countries have strategies focused at the producer part of the supply chain. Australia, New Zealand, and the United States also emphasized support services. Within support services (64%), Australia noted the need for increased warehouse capacity and transportation coordination regionally in a disaster. It also focused actions on distributors (58%), such as food service and retail stores that are critical in providing food for disrupted communities during natural disasters. New Zealand also emphasized support services (31%), due to the vulnerability of roads, rail, ports, and airports to climate change and natural hazards. The United States also gave emphasis to support services (40%), inclusive of all forms of food transportation across the United States. Sweden chose to focus more heavily on food distribution (37%) to consumers but by providing strategic guidance on the alignment of producers and processors to providing the sustainably produced, organic, and healthy foods that are being demanded by consumers, including tourists to Sweden.

Figure 2c shows a comparison of the shocks and stressors targeted by countries’ actions. When addressing food system vulnerabilities to different shocks and stressors, most countries addressed the stressor of climate change, except for Australia, which focused on natural shocks (100%), floods and bush (forest) fires. Given the timing of its plan’s development, the US also placed greater emphasis on addressing biosecurity hazards (40%) and pandemic vulnerabilities (33%) due to the COVID-19 pandemic. New Zealand’s primary focus was on climate change resilience (91%), since the documentation analyzed was climate related. The other category (26%) was infrastructure, which was outlined by the US in terms of failing infrastructure that requires maintenance or updating as it is outdated.

Figure 2d provides a comparison of the implementation level of actions for the four countries in this study. All included countries centered most of their actions at the national level. This would be expected since we reviewed national plans. Australia did, however, emphasize coordination between national and local levels (61%). It is notable that New Zealand also included a local (33%) and community (32%) emphasis, due to climate adaption planning requiring coordination between national and local government authorities, as well as a focus on climate risk prone communities (such as Māori and rural communities). Sweden’s notably few actions outside of the national level was due to the objective of setting strategic direction at the national level.

Figure 2e shows the intended effect on food security of the food system resilience actions. We found that, amongst all countries, increasing food availability and food sustainability was the most expected and targeted outcome of the actions. Food system sustainability and food system resilience are not synonymous: food system sustainability is the ability of the food system to meet the needs of future generations and is a component or part of food security and food system resilience. Australia (87%), Sweden (68%) and the United States (61%) placed more emphasis on availability. Sustainability was addressed by the United States (70%) and New Zealand (57%) as they sought actions to increase resilience in the face of climate change. Access was addressed to a lesser extent by New Zealand (17%) and Sweden (11%), although still more than the other countries, due to their focus on aligning producers and processors with their consumers’ needs. Agency was also addressed by New Zealand (17%) as it sought some actions to address inequities that they acknowledged in the face of climate change.

### Food System Resilience Areas of Policy Focus

In the next section, we explore the focal policy areas where countries concentrate on the same combination of categories in their policy actions. Table 3 lists the repetitions of category combinations in a country’s resilience policy actions. In these instances, we found more than one action targeting the same combination of resilience attributes, parts of the supply chain, shocks and stressors, implementation level, and intended effect of food security. We found that governments’ policy actions did repeat across some combinations of categories, however not significantly.

**Table 3:** Policy focus areas with two or more actions targeting the same combination of categories.

| Country | Number of Policy Action | Food System Resilience | Food System | Shocks & Stressors | Implementation Level | Food Security |
| --- | --- | --- | --- | --- | --- | --- |

|  | **Repetitions** | **Attributes** | **Supply Chain** |  |  |  |
| --- | --- | --- | --- | --- | --- | --- |
| **US** | 3 | Adaption | Input Services | Climate | National | Sustainability |
| **NZ** | 2 | Awareness | Support Services | Biosecurity | National | Sustainability |
| **NZ** | 2 | Equity | Producer | Climate | National | Agency |
| **Sweden** | 2 | Diversity | Producer | All hazards | National | Availability |
| **Sweden** | 2 | Redundancy | Producer | Climate | National | Sustainability<br>Availability |
| **US** | 2 | Awareness<br>Adaption | Input Services | Climate | National | Sustainability |
| **US** | 2 | Awareness | Producer | Climate | National | Availability |
| **US** | 2 | Diversity | Support Services | Epidemic or Pandemic | National | Availability |
| **US** | 2 | Adaption<br>Capital<br>Reserves<br>Other | Producer | Climate | National | Sustainability |
| **US** | 2 | Adaption | Producer | Climate | National | Sustainability |
| **US** | 2 | Equity<br>Capital<br>Reserves<br>Other | Producer | All hazards | National<br>Community | Availability<br>Agency |
| **US** | 2 | Preparedness | Producer | Biosecurity | National | Stability |

We found that, of the 167 possible policy combinations, in only 12 cases did the same combination occur 2 or more times. The United States had 67 unique combinations and only 8 repeating combinations. The United States’ top combination (row 1) indicates a repetition of national climate change policy actions directed at adaption of input services. New Zealand had 52 unique combinations with only 2 repetitions. New Zealand’s top combinations, in rows 2 and 3 (Table 3), show that, at a national level, there is a focus on developing support services for biosecurity to maintain the food system in the future and to establish equity for Māori producers who are projected to suffer disproportionate disruption in their indigenous food system as a result of climate change. Australia had 31 unique combinations and no repetitions. Sweden had 17 unique combinations with only 2 repetitions. This indicates that governments are using a variety of actions to address food system resilience issues within their countries.

We also analyzed the temporal aspects of the countries’ actions and found that they were primarily focused on short- and medium-term actions, with 84 short-term and 57 medium-term actions, and only 7 slated for the longer term. Governments looked to the past and future to inform actions, with 137 actions being based on retrospective reviews and 134 taking a prospective viewpoint, while 39 considered both prospective and retrospective viewpoints.

## Discussion

Comparing the identified national food system resilience documents of Australia, New Zealand, Sweden, and the United States, we found that countries are developing approaches using a variety of resilience attributes, target different parts of the food supply chain, address a range of shocks and stressors, focus at different scales, and seek to have an effect on food security. When comparing within countries, to address the same issue a country may at times utilize multiple actions using an identical combination of categories but, on closer examination, these actions are comprised of multiple and differing policies and investment levers, even though their categorization is the same.

The analysis framework is useful for highlighting gaps and identifying government focal areas as they address food system resilience within their countries. This framework can also be used to develop recommendations for countries: supplementary information file 4 provides an example for New Zealand. Comparing across countries using the framework can assist in determining the expected results of different approaches and can also be used to monitor the results of policy actions and their intended effects on food security, for improved evidence-based policymaking and refinement over time.

From our analysis, we identified several potential gaps where there were fewer actions. One identified gap was that there are relatively few actions across all countries that address the resilience attributes of capital reserves (financial, social, and natural) or equity (procedural, distributional, structural, and intergenerational). Capital reserves are useful as they are resources that are set aside for use during shocks and/or stressors. Addressing inequities is useful in preparation for, responding to, and recovering from shocks and/or stressors to eliminate unequal outcomes for certain populations. Countries have focused on building reserves to support needs that include but also extend beyond the food system. For example, the United States created a federal stockpile of personal protective equipment and vaccines for food system workers because of the COVID-19 pandemic, while in New Zealand there was discussion of whether there was sufficient petroleum in or readily available for the national reserve, petroleum being critical at present for food production and distribution. Addressing equity in the food system to promote resilience and food security is another gap that requires further attention by governments, New Zealand and the United States have taken some equity related actions, yet food system equity and food security still remain of national concern and require further focused actions. Policymaking and food system resilience planning efforts that proactively consider procedural, distributional, structural, and intergenerational equity can help to build food systems that are more equitable and just, even if a disaster never occurs. Our analysis also found additional relative gaps across countries in the attributes of connectivity, diversity, and redundancy, and thus further action may still be required on the part of governments to support the development of those attributes within their country’s food system.

Another key gap was in the types of disasters considered by countries. Aside from a brief mention due to the China-United States trade conflict’s being an issue in the United States, economic and political crises were not explicitly considered except in the case of Sweden. Sweden mentioned that, over the course of its membership in the European Union (EU), the country has developed an over-reliance on food imported from other EU countries, which has led to a resurgence of issues with food security in Sweden. Australia and New Zealand’s economies rely on trade of their food products, and therefore it is important to consider modeling political and economic crisis impacts on the food system, and moving from being tactical to being more strategic, prepared, and resilient. The food system is becoming increasingly reliant on digital food supply chains’ logistical systems, and therefore cybersecurity is becoming an area for consideration by governments in food system resilience planning. Australia amended its critical infrastructure bill to include food companies, resulting in obligations to report cybersecurity attacks.

Based on our analysis governments emphasized improving food availability. There was little emphasis on the other food security sub-categories: access, agency, utilization and acceptability, and stability in food security. Food access is important for policymakers to consider, given the significant threat that any shock and stressor will challenge economic and physical access, such as high food prices or decreases in net income, as well as populations that may not have the ability to physically access food stores. Enhancing food system utilization and acceptability for the long term, and preventing intergenerational food insecurity, are known to support well-being. These can also be important considerations in the recovery cycle from an emergency: Australia noted it could take up to 6 months for the food system to be restored. Given the compound and long-term stressors on food systems, countries can now be in a situation of having to face deepening intergenerational food insecurity. In addition, countries are considered food insecure if the food available is not acceptable, dignifying, and culturally appropriate. NZ’s government has publicly committed to work with the Māori community on climate change planning and acknowledges their traditional coastal food sourcing. This commitment should, however, be broadened to include other shocks and communities. There was also an absence of actions that create stability from disruption in the current food system, with more actions taken to support future food systems than to address current food system needs.

We identified four key emergent themes when comparing countries: lack of competition in the food system, diminishing water quality and quantity, new climate-resilient pests and invasive species, and transportation bottlenecks (“chokepoints”). New Zealand, Sweden, and the United States all stated that there were vulnerabilities emerging within their food systems due to a lack of competition, highlighting consolidations in parts of their supply chain, over-reliance on certain food products and markets, and rules and regulations that hinder competition and sustainable production. Water quantity in the food system was another important aspect that the four countries highlighted. The United States and New Zealand are projecting droughts and water shortages in production, while Australia, by contrast, states that water is a key dependency across the entire food supply chain, not just production. The United States, New Zealand, and Sweden are actively working on the development of advanced decision-making tools for water resource planning and management to support producers and local resource management authorities. Those countries are all planning to increase surveillance of their food systems by providing additional resources to their national laboratories and quarantining facilities. These measures aim to prevent the introduction of exotic new pests and diseases, and other invasive species expected as a result of climate change and global trade and movements. Transportation systems allow food to move from farms to tables or to borders, ports, and airports, utilizing different combinations of transportation depending upon the producer’s location and the type of food. Countries are actively highlighting the risks to the transportation of food in emergency situations, such as significant concerns about the risk of aging food transportation infrastructure and the need for investment, modernization, increased capacity, and diversity at transportation system chokepoints. Others are highlighting the need to address resilience, reliability, and preparedness for potential disruptions—including climate change—across existing, modified, and new transport infrastructure, highlighting the need in emergency situations for flexibility, diversity, redundancy, and coordination across the food transportation system.

The study has several limitations. It is important to note that the countries included all have extensive activities aimed at strengthening their food systems and that, while those activities may also have an effect on building resilience, they are not always framed as resilience or directly addressing shocks or stressors. Nonetheless, the studied documents reflect an important component of a country’s food system resilience work, and were collated and analyzed based on the method outlined. In addition, as New Zealand has not yet developed a food system resilience government review, strategy, or policy, we used climate change-related documentation as a proxy. Future studies could include climate change documentation and additional food system documentation, and a New Zealand-wide food system resilience plan once developed, to draw out further parallels and differences between countries. The four countries included in this analysis are not an exhaustive list of countries that have published national food system resilience documents. The countries were determined based on the selection criteria, and future work could include low- and middle-income countries. While coding was conducted based on the definitions outlined, coding is subjective, although reviews helped to reduce differences between coders. It is also important to note that each country has its own unique context and that perhaps some concentrations or gaps could be appropriate for that country (for example: Sweden highlighted the issue of food insecurity and its strategic focus on structural action to increase redundancy and diversity in the food system). More extensive research would be needed into each country’s context to understand the approach taken, the prioritization process, and ultimately the food security outcomes. Finally, the study encompassed issues and actions directed within a country; international aspects of the food system were mentioned in brief but were not considered in the analysis. This could be an important component to include for future research: how countries are impacted by—and impact—global trade and food system resilience.

Despite these limitations, this study provides a cogent framework that maintains the integrity and the broadness inherent to the theory of food system resilience. The framework is designed for a national-level analysis, unlike other frameworks that are limited to a part of the food system, such as a city or producers. It also provides a mechanism to monitor and evaluate food system resilience planning and food security outcomes.

## Conclusions

This comparative analysis of national-level food system resilience activities finds that work has been done towards preparing for climate change and other natural disasters, although not as much has been done for other shocks or stressors. Countries are utilizing multi-pronged policy actions to address food system resilience issues, and are focused on both retrospective reviews and prospective models of disruptive events to inform their planning.

This work supports policymakers and academics by distilling what is covered by the selected governments pursuing food system resilience approaches, including commonalities and potential gaps, and also by synthesizing the actions already undertaken and identified as resilience-focused. Through categorizing food systems resilience actions, we can start to distill the complexity of government policy actions to address food system resilience, and provide insights into the emphasis, focal areas, and themes in current food system resilience work by governments. This framework may also be useful to different levels of government by providing a method for assessing how their policy actions support different components of food system resilience.

## Data Availability

The dataset used and analyzed during the current study are available from the corresponding author on reasonable request.

## Declarations

Ethics approval and consent to participate Not applicable.

## Consent for publication

Not applicable.

## Authors’ contributions

JL conceived and carried out the study, performed data coding, co-developed the data analysis method and approved the results, and drafted the manuscript. EM reviewed the conceptual framework for the study, reviewed data coding, agreed coding definitions, and edited and reviewed the manuscript for publication. LD developed the data analysis and R code, and reviewed the methods and results section related to the analysis. RN reviewed and approved the conceptual framework of the study and reviewed the manuscript for publication. All authors read and approved the final manuscript.

### Acknowledgements

David Lindsay provided insightful review and edits of the draft manuscript.

## Authors’ information

JL earned her Master of Public Health from Johns Hopkins Bloomberg School of Public Health, a Master of Arts from City College, and a Bachelor of Consumer and Applied Sciences from Otago University, NZ. She has over two decades of experience working for leading food organizations across three continents, and in the environmental sector developing knowledge products to inform on environmental and social risk in food supply chains.

EM is a Ph.D. candidate in the Department of Environmental Health and Engineering at the Johns Hopkins Bloomberg School of Public Health. She has worked as a research assistant for Center for a Livable Future on an urban food system resilience project with five cities across the US. Before starting her Ph.D., Elsie worked on several projects related to the United Nations Sustainable Development Goals. Elsie earned her Master of Public Health in Social and Behavioral Sciences from the Yale School of Public Health and a Bachelor’s in Diplomacy and World Affairs from Occidental College.

LD earned her Master of Science (Integrated Studies in Land and Food Systems) from the University of British Columbia and a Bachelor of Arts (Environmental Sciences and German Studies) from the University of Virginia. She is a Sustainability Scientist in Agriculture and Sustainable Food Systems at The Nature Conservancy.

RN has a PhD from Johns Hopkins Bloomberg School of Public Health, an ScM from Harvard School of Public Health, and an AB from Brown University. She is an Associate Professor at the Department of Environmental Health and Engineering, Johns Hopkins Bloomberg School of Public Health and co-leads the department’s PhD track in Environmental Sustainability, Resilience and Health. Her research, policy and practice activities focus on advancing change at the intersection of food systems, planetary health and social justice. She also works with public health practice and with broader food system issues through her longtime role at the school’s Center for a Livable Future, an academic center focused on food systems & public health.

