## Supplementary Information File 1 for "A comparative national-level analysis of government food system resilience activities in preparation for future food system disruptions"

**Data collection categories, sub-categories, and definitions**

**Food System Resilience Attributes**

Refers to characteristics of the food system known to increase resilience.

- Adaption [1]

The system is flexible and can adapt to changing circumstances, modifying behaviors and adapting existing resources to new purposes. Policies that encourage adaption within a system include ones that promote new leader training [2], support local business development, and facilitate efficient information flow between academic, private, and government sectors.

- Awareness [1]

The food system has knowledge of its assets, liabilities, and vulnerabilities. This includes situational awareness, which allows for assessing new information and adjusting to shocks and stressors in real time. Policies that promote awareness within a system include funding research or disseminating information about assets, liabilities, vulnerabilities, and monitoring.

- Capital Reserves [3]

Having social, financial, natural, political, food, and food input and supply reserve “backup” resources that can be used during a disruptive event for example: strong community networks (social), reserve funds (financial), arable soil (natural), state government support (political).

- Connectivity [1,4]

Policies that promote integration and coordination among food system components such as transportation and information technology to establish dynamic information streams between different governance levels within the system.

- Diversity [3,4]

Having a variety of food system elements that can serve a similar purpose for example: a variety of retail options for food such as farmers markets, independent grocers, and supermarkets.

- Equity [3]

Is inclusive of the following categories of equity and inclusion in the food system:

- Procedural equity: having “transparent, fair, and inclusive” food system resilience planning, implementation, and evaluation processes.
- Distributional equity: having the benefits and burdens of food system resilience planning that work to reduce inequities.
- Structural equity: having resilience actions uproot long-term embedded structures that perpetuate inequitable food system and resilience outcomes.
- Intergenerational equity: having actions taken today conserve resources for future generations.
- Redundancy [3,4]

Having multiple or duplicative food system elements that can serve the same purpose for example: fertilizer sources with more than one country providing inputs.

- Preparedness [3]

Having a plan in place for how to ensure food access, availability, acceptability, and agency during a disruptive event for example: food included in emergency management protocol.

**Food System Supply Chain**

Refers to the broad categories below that were used as they represent most of the components of the food supply chain, with some exceptions based on the Food and Agriculture Organization (FAO) [4] definitions.

- Producer: the producer category includes food from agricultural and horticultural origins. Agriculture and horticulture here mean all subsectors: crops, livestock, pastoralism, fruit, vegetables, nuts, fisheries, and aquaculture. Food may be produced by large-scale producers and business enterprises, and by small-scale producers and small and medium food enterprises (SMEs).
- Processor: a food processor means a food establishment that processes, manufactures, wholesales, packages, or labels, food such as meat, or engages in fruit packing, cheese making or consumer goods manufacturing etc.
- Distributor: refers to a food retailer (such as supermarkets, co-operatives, farmers markets etc.) or food service provider (such as cafes, restaurants, hospitals, and schools etc.) that have physical or virtual stores that provide food to consumers.
- Input Services: provide variable inputs, such as seed, fertilizer, fuel and labor, and quasi-fixed inputs, such as farm machinery, milling machines and coolers for perishables.
- Support Services: include actors and activities for movement of inputs, outputs, and factors such as transport and storage operators, connecting production to consumption.

**Shocks and Stressors**

Refers to the type of shock or stressor being addressed by the action.

- Biosecurity: refers to harmful pests and diseases that can cause damage to plants and animals.
- Climate: refers to the stressor of climate change that has a multiplying effect to other stressors or shocks. It includes the effects of sea level rise, increased temperatures, coastal erosion, and more frequent extreme weather events.
- Cybersecurity: refers to shocks to digital technologies by exploited controls and practices to gain initial access or as part of other tactics to compromise cyber systems.
- Economic & Political Crisis: refers to a shock that is economic and political in nature (domestic or international in origin) that can have an unexpected large-scale impact on the economy.
- Epidemic or Pandemic: refers to a human disease outbreak that, in the case of an epidemic, has an unexpected increase in the number of disease cases in a specific geographical area and that, in the case of a pandemic, exhibits disease growth that is exponential and covers a wide area, affecting several countries and populations.
- Natural: refers to shocks or disasters that occur naturally, such as earthquakes, tsunamis, hurricanes/cyclones, tornados, landslides, floods, and droughts.

When actions refer to all hazards, or a “crisis” that is undefined, all shocks and stressors were selected.

**Implementation Level**

Refers to where the action is directed for implementation.

- National: at the country or central government level
- Regional: at the regional or multi-state or multi-province level
- Local: at the state or provincial level of government
- Communities: at the social group level who share a geographic location, culture, or heritage
- Household: at the familial or extended family level

**Food Security**

Refers to the latest food security enhancing categories outlined in a report on Food Security and Nutrition by a high-level panel of experts [5].

- Access: policies that make healthy food more financially and physically accessible including addressing uneven quality of food environments, distribution, and access to markets for small-scale producers; increasing retailer diversity, and; reducing the distance between production and consumption.
- Agency: policies that consider an individual’s right to food, fair and equal consideration of communities that affect the food system, equal access to information and technology about the food system.
- Availability: policies that increase the amount of food in the food system by: increasing yields; increased investment in agriculture and horticulture; efficiencies in production; reduction in post- harvest losses in handling and transport, and retail food waste, and; ensuring seasonal workforce availability and reduction in the effect of shocks and stressors on production.
- Stability: policies and planning that reduce instability or variability in the current food system from causes such as pests, food safety issues, economic crises, and trade disruptions that create volatility in food prices and food input prices as shocks and stressors to the food system.
- Sustainability: policies that reduce the impacts to the future food system from causes such as new pests, new diseases, degradation of natural resources for future generations; climate change affecting future production; biodiversity loss damaging genetic diversity; resource inefficiencies and pollution from overuse of agrochemicals or concentrate of animal waste; ecological and economic costs of unsustainable agriculture, unsustainable diet, and an aging farming population.
- Utilization and Acceptability: policies that ensure food that is safe, acceptable, and culturally appropriate, and provides sufficient nutrients and micronutrients to maintain good health for example: increasing diversity in the diet, nudges to healthy and sustainable diets, addressing the urban environment including food environments and obesity.

**Timelines**

Refers to the timeline to define when the action would be completed based on the definition in the documentation.

- Short term: up to 2030, or continuing and existing action or funding;
- medium term: 2030 to 2050;
- long term: 2050 to 2100.

When no timeframe is given for an action, it was left blank, or if it was not defined in years but as either short, medium or long term, then those categorizations were used.

**Perspective**

Refers to whether the view of the issue and/or action was based on the past or future shocks or stressors**.**

- Retrospective: refers to policy Issues and actions based on previous issues and vulnerabilities experienced for example: a previous flood.
- Prospective: refers to policy issues and actions based on projections of future vulnerabilities for example: climate change projections.
