## Supplementary Information File 2 for "A comparative national-level analysis of government food system resilience activities in preparation for future food system disruptions"

**R Code for Analysis**

################################################################################

\# Comparing food resilience response actions by country according to response

\# coding in Lloyd et al. (2023).

\#

\# This code is intended to analyze individual country actions across the

\# resilience categories

\#

\# Code prepared by Lyndsey Dowell for research conducted in Lloyd et al. (2023)

\# Last Updated: January 2023

################################################################################

#install.pakages("dplyr")

library(dplyr)

\# Set data directory

\# Read in issue and action data

raw <- read.csv("FSR_Coding.csv")

\# Replace NAs with 0 - recommended step

\# raw <- raw %>%

\# mutate_at(c(4:47), ~replace(., is.na(.), 0))

\# Read in categorization for issue and action coding

cat <- read.csv("cat_ids.csv")

\# Subset action data for analysis

cat_list <- unique(cat[,1])

data_list <- list()

\# Loop through to create data frames for each analysis category

for(i in 1:length(cat_list)){

df <- raw[,c(1,4)]

c = 2

for(j in 1:nrow(cat)){

if(cat[j,1] == cat_list[i]){

c = c + 1

df <- cbind(df,raw[,which(colnames(raw) == cat[j,2])])

colnames(df)[c] <- as.character(cat[j,2])

}

}

data_list[[length(data_list)+1]] <- df

names(data_list)[i] <- as.character(cat_list[i])

}

\# Function to create actions pivot table

pivot <- function(data){

table <- data %>%

filter(ACTION == 1) %>% # Only looking at actions

group_by(COUNTRY) %>% # Grouping results by country

summarize_at(c(1:(ncol(data)-1)),sum, na.rm = TRUE) # Calculating action frequency

}

\# Create and export pivot tables for each analysis category

for(i in 1: length(data_list)){

pv <- pivot(data_list[[i]])

write.csv(pv, paste(as.character(cat_list[[i]]), 'pivot', 'csv', sep = '.'))

}

################################################################################

\# Looking for repeated action attribute combinations across and within countries

\# (Lloyd et al., 2023).

\#

\# This code is intended to count the repetitions of identical attribute

\# combinations across all resilience categories

\#

\# Code prepared by Lyndsey Dowell for research conducted in Lloyd et al.(2023)

\# Last Updated: January 2023

################################################################################

#install.packages("dplyr")

#install.packages("tidyr")

library(dplyr)

library(tidyr)

\# Set data directory

\# Read in issue and action data

df <- read.csv("FSR_Coding.csv")

\# Narrow list down to just actions

df2 <- df %>% drop_na(ACTION)

\# Look for combinations of a subset of attributes

\# Combination set 1: look at frequency of identical action coding across all

\# countries to identify where countries have the same responses

\## (remove country, policy descriptors, time frame, and other descriptors)

combo_all <- df2[,c(5:13,15:25,26:32,34:39)] %>% #sub-setting to desired attributes

group_by_all() %>%

summarise(count = n()) %>%

arrange(desc(count))

\# Assign combination ID

combo_sub1 <- combo_sub1 %>%

mutate(ID = row_number())

\# Combination set 2: include country in subset to see frequencies of identical

\# action coding specific to individual countries

\## (remove policy descriptors, time frame, and other descriptors)

combo_by_country <- df2[,c(1,5:13,15:25,26:32,34:39)] %>% #sub-setting to desired attributes

group_by_all() %>%

summarise(count = n()) %>%

arrange(desc(count))

\## Export results tables

write.csv(combo_all, "attribute_combinations_altogether.csv")

write.csv(combo_by_country, "attribute_combinations_individual.csv")
