## Supplementary Information File 4 for "A comparative national-level analysis of government food system resilience activities in preparation for future food system disruptions"

**The Case of New Zealand’s Food System**

New Zealand (Aotearoa) is an island nation in the South Pacific Ocean formed on the collusion zone (fault line) between the Pacific and Australia tectonic plates, creating a high risk of earthquakes, volcanic activity, and tsunamis [1]. New Zealand is experiencing the effects of climate change, such as sea levels rising, and extreme weather events that are more frequent and severe. The National Institute of Water and Atmospheric Research (NIWA) [2] projects that changes in temperature and seasonality will have implications for agriculture and horticulture, and affect where certain crops can be grown. Droughts are projected to increase in frequency and severity, making New Zealand’s agriculture sector vulnerable to declining crop yields and pasture growth. Projected changes in ocean temperature, chemistry, and currents will have impacts on New Zealand’s marine life, fisheries, and aquaculture [1].

In the medium term, the Climate Change Risk Assessment Report [2] warns of supply chain and distribution network disruptions due to extreme weather events. New Zealand is also geographically separated from its main trading markets and is economically dependent upon international trade for its primary products, which include agriculture, forestry, fisheries, and aquaculture [3] (5.7% GDP) [4], making it particularly prone to supply and distribution disruptions. New Zealand has a growing food insecurity issue. In 2020, it was reported that 14.5% [5] of the population was moderately or severely food insecure.

We have therefore decided to undertake a comparative analysis of food system resilience policies at a national level to determine the usefulness of such an analysis in informing policy development, in this case for New Zealand due to its risk profile.

**Recommendations for New Zealand**

Given the vulnerability and sensitivity of New Zealand to all shocks and stressors, and the significant economic and cultural heritage embedded into its food system, as well as the increasing failures within the food system to protect the populace from food insecurity and non-communicable diseases (associated with diet), the research shows that New Zealand’s recent climate policy agenda has begun to indirectly address a number of food system resilience related issues.

Based on the analysis, New Zealand could go further and benefit from considering a review of the following regarding its food system resilience: diversity, redundancy, connectivity, capital reserves, while continuing to work on equity strategies that support processors, distributors, and food system support services at the regional, local, community, and household levels to addressing access, stability, and utilization in food insecure areas.

**Addressing Diversity**

Diversity is the key to building the capacity to absorb shocks in the food system, such as including diversity in food sources and a diversity of actors in the food system. Absorptive capacity is determined by the food system’s structural characteristics, such as the number of diverse stakeholders involved, the institutions that coordinate them, and the infrastructural robustness that they rely on [6]. On a dietary sourcing flexibility index (DSFI) that FAO [6] developed, New Zealand was lower compared with the US, Australia, and Sweden. Therefore, domestic production, stocks, and imports were less likely to make up a healthy diet for the population. Possible policy strategies could be:

- Supporting diversity in domestic production of fruits, vegetables, and other perishables for availability, access, and utilization by the food retail sector, ensuring food security, nutrition, and health.
- Supporting diversity and accessibility to food: school lunches, food banks and pantries, and emergency food social safety nets.
- Reviewing the diversity of food storage, processing and value-added facilities, their spatial distribution, and diversity in transportation routes to distributors.
- Supporting diversity and access to exported and imported products and markets, to absorb supply and demand shocks.
- Supporting diversity in food system infrastructure, including the ability to switch, as well as having multiple sources of clean water and electricity to large, small, and medium food processors, support services, and distributors.
- Ensuring sufficient diversity and availability of nutritionally rich foods, and year-round access.

**Redundancy**

Redundancy is the duplication of critical components or functions of a food system [6], including back-up systems, processes, or stores. Examples of this might include spare inventory, alternative transport routes, and strategic food stocks at the national level. Incorporating redundancy into the food system requires a balance between costs, efficiency losses, and resilience benefits [6]. When assigning policy priorities, it is important to consider that SMEs face financial and logistical constraints to investment in redundancy. The challenge is to implement the right mix of policies and interventions that help build capacities, minimize trade-offs, and create synergies that lead not just to resilience but also efficient and inclusive food supply chains [6]. Policy options might include:

- Provide guidance to supply chain organizations to review dependencies and encourage sourcing redundant and/or substitutable processing components or inputs for business continuity.
- Support service facilities to develop multiple storage and delivery options including surge capacity. This could include developing partnerships with other, complementary companies too.

**Connectivity**

Connectivity contributes to absorptive capacity by providing options once a disruptive event happens. The food system is comprised of interconnected activities performed by various actors. Farmers, processors, and distributors depend on actors in lateral chains to supply inputs, and logistics and transport services. All are vulnerable to diverse risks, shocks, and stressors, whose impacts can disrupt food systems [6].

- Therefore, the interconnection of food system actors means that resilience depends on the decisions and overall performance of actors in the food system. The decisions made by one actor can have implications for the others up and down the food system. Therefore, all actors from the public and private sector should, first, assess the degree of diversity and connectivity in their food distribution network. Second, they should examine the availability of alternative routes (redundancy) when transportations links are severed. Third, they should assess the relative cost of a variety of detours, and the impact of disruptions.
- The government plays a key role in creating an enabling environment for improved coordination in food supply chains, for example, through public investments in infrastructure, emergency management coordination and training, and resource management. This type of investment can in turn support the incorporation of food access into regional and municipality planning of infrastructure and transportation into urban centers for distributors (food retailer and food service).
- There is also an important role for policymakers to play in connecting and aligning nutrition policies with food system and public health policies, and ensuring that nutritional needs are aligned with production.

**Equity**

Addressing equity and systemic injustice in food system resilience development requires that the community is fully engaged before, during, and after a disruption, and the consideration that not everyone’s needs for support are the same, and ensuring that the differential needs are addressed.

- Procedural equity

Begin food system resilience planning with the people and communities most likely to be impacted by a disruption, establishing an open dialogue and co-development of the steps to build a more resilient food system [7].

For example: this includes ensuring that the community is representative of all the diverse members and viewpoints present in the community.

- Distributional equity

Distributional equity means resources are prioritized to communities with the greatest vulnerability and inequalities [7]. The causes and impacts of food system disruptions, and the resources available to recover from them, are not equally distributed across all communities. Low-income and marginalized communities, including Māori, Pacific Islanders, and other immigrants are more likely to have fewer economic resources to prepare for and overcome a crisis [7].

For instance: this includes equal access to emergency food and water supplies, to storage, and access to cooking tools and skills at the household level.

- Structural equity

Structural equity means the goal of the food system resilience plan is not to maintain the status quo but to create a food system that is more equitable, by looking to the reasons that the food-related health inequities were caused, and addressing them in planning [7].

For instance: equitable access to land and production resources, or ensuring that the crops grown and available reflect the culturally diverse needs of the community or region.

- Intergenerational equity

Intergenerational equity means that food system resilience planning considers the effect of actions taken today on long term access to food system resources.

For instance: a new climate smart road being planned through a Māori community.

**Conduct a nationwide food system all hazards vulnerability assessment:**

- Develop models and map all hazards and timelines across the country, including retrospective scenarios (policy planning for a similar shock) but also prospective scenarios across a wide variety of shock scenarios, and how they could destabilize various aspects of the food system [8]. It is also important to consider how current trends in the food system mitigate or exacerbate these scenarios [8]. It would therefore be important to include retrospective research from previous natural and biosecurity shocks, and the COVID-19 pandemic, as well as prospective modelling for all other hazards outside of climate change.
- Develop maps of all rural, peri-urban, and urban production areas, key transportation routes, processing facilities, warehousing (storage facilities), and distribution sites that are destined for both domestic and export food markets.
- Develop maps of all institutions for vulnerable people (young, elderly, disabled, incarcerated, healthcare facilities); areas of low socio-economic status; areas of food insecurity, and; food retailers and food service outlets and their food transport and distribution routes [5,9].
- Develop a food system resilience program with communities and households in the areas that are most vulnerable and likely to be impacted to shocks and stressors [7]:
  i. Develop a fault tree diagram [5] for the community’s food systems and what would what it is means to have their food system working well, and what would they define as a food system failure?
  ii. Determine with the community critical food system assets to ensure food security.
  iii. Map these community critical food system assets and their connections to local, regional, and national food system assets and transportation infrastructure.
- Explore different models’ food system dynamics and multiple time scales, and determine scenarios for policy makers, supply chain stakeholders and communities that highlight that the food system is close to failure [10].
- Embed food systems into emergency management and critical lifelines and infrastructure at national, local, and community levels, and provide training and expertise on food systems.
- Determine priorities and timelines to maximize impact based on resources available.
- Based on the quantitative and qualitative data elicited from the above and other sources, determine recommendations for national food system resilience.

The above recommendations are a starting point based on the current analysis. To review and prepare for future disruptions to New Zealand’s food system, additional data would need to be identified and analyzed, and collaboration would be needed on food system resilience between New Zealand's national and local government authorities, and stakeholder communities, with the objective of creating a plan, leveraging government planning guides such as the Johns Hopkins Center for A Livable Future’s Food System Resilience: A Planning Guide for local governments [7].
